# Personalized Connectome Mapping to Guide Targeted Therapy and Promote Recovery of Consciousness in the Intensive Care Unit

**DOI:** 10.1101/19013987

**Authors:** Brian L. Edlow, Megan E. Barra, David W. Zhou, Samuel B. Snider, Zachary D. Threlkeld, John E. Kirsch, Suk-tak Chan, Steven L. Meisler, Thomas P. Bleck, Joseph J. Fins, Joseph T. Giacino, Leigh R. Hochberg, Ken Solt, Emery N. Brown, Yelena G. Bodien

**Affiliations:** Center for Neurotechnology and Neurorecovery, Department of Neurology, Massachusetts General Hospital, Boston, MA; Athinoula A. Martinos Center for Biomedical Imaging, Massachusetts General Hospital, Charlestown, MA; Department of Pharmacy, Massachusetts General Hospital, Boston, MA; Department of Brain and Cognitive Sciences, Massachusetts Institute of Technology, Cambridge, MA; Department of Neurology and Neurological Sciences, Stanford School of Medicine, Stanford, CA; Department of Neurology, Northwestern University Feinberg School of Medicine, Chicago, IL; Division of Medical Ethics and Consortium for the Advanced Study of Brain Injury (CASBI), Weill Cornell Medical College, New York, NY; The Rockefeller University; Solomon Center for Health Law and Policy, Yale Law School, New Haven, CT; Department of Physical Medicine and Rehabilitation, Spaulding Rehabilitation Hospital, Boston, MA; School of Engineering and Carney Institute for Brain Science, Brown University, Providence, RI; Veterans Affairs RR&D Center for Neurorestoration and Neurotechnology, VA Medical Center, Providence, RI; Department of Anesthesiology, Massachusetts General Hospital, Boston, MA

**Author notes:** **Corresponding author:** B.L. Edlow. **Study Funding:** The study was funded by the NIH Director’s Office (DP2HD101400), National Institute of Neurological Disorders and Stroke (K23NS094538, R21NS109627, RF1NS115268), American Academy of Neurology/American Brain Foundation, James S. McDonnell Foundation, Rappaport Foundation, and Tiny Blue Dot Foundation. We also acknowledge support from the National Institute of Neurological Disorders and Stroke Clinical Trial Methodology Course (R25NS088248). This work was conducted with support from Harvard Catalyst | The Harvard Clinical and Translational Science Center (National Center for Advancing Translational Sciences, National Institutes of Health Award UL 1TR002541) and financial contributions from Harvard University and its affiliated academic healthcare centers. The content is solely the responsibility of the authors and does not necessarily represent the official views of Harvard Catalyst, Harvard University and its affiliated academic healthcare centers, or the National Institutes of Health.

**Keywords:** coma, consciousness, brain injury, connectome, clinical trial, biomarker

## Abstract

There are currently no therapies proven to promote early recovery of consciousness in patients with severe brain injuries in the intensive care unit (ICU). Early recovery of consciousness would benefit patients and families by reducing the likelihood of premature withdrawal of life-sustaining therapy and may decrease ICU complications related to immobility, facilitate self-expression, enable autonomous decision-making, and increase access to rehabilitative care. Here, we present the connectome-based clinical trial platform (CCTP), a new mechanistic paradigm for developing and testing targeted therapies that promote early recovery of consciousness in the ICU. The scientific premise of the CCTP is that personalized brain connectome maps can be used to select patients for targeted therapies that promote recovery of consciousness. Structural and functional MRI connectome maps will identify circuits that may be amenable to neuromodulation. Patients will be selected for clinical trials in the CCTP paradigm based on connectomes that are likely to respond to targeted therapies. To demonstrate the utility of this precision approach, we describe STIMPACT (<u>S</u>timulant <u>T</u>herapy Targeted to Individualized Connectivity <u>M</u>aps to <u>P</u>romote Re<u>ACT</u>ivation of Consciousness), a CCTP-based clinical trial in which intravenous methylphenidate will be used to promote early recovery of consciousness in the ICU (ClinicalTrials.gov NCT03814356). We propose that the CCTP has the potential to transform the therapeutic landscape in the ICU and improve outcomes for patients with severe brain injuries.

## Introduction

For patients with severe brain injuries in the intensive care unit (ICU), the range of possible outcomes includes death, a persistent disorder of consciousness (DoC), recovery of consciousness with functional disability, and recovery of functional independence [1]. Early recovery of consciousness in the ICU is a strong predictor of long-term outcome [2-4] and a critical determinant of decisions to continue or withdraw life-sustaining therapy [5]. However, there are currently no treatments proven to promote early recovery of consciousness in the ICU. Without knowing whether a patient can recover consciousness, many families – guided by intensive care clinicians – withdraw life-sustaining therapy, a decision that accounts for up to 70% of deaths in ICU patients with severe traumatic brain injury (TBI) [6, 7] and over 40% of deaths in patients with hypoxic-ischemic injury [8]. Furthermore, decisions to withdraw life-sustaining therapy often occur in the first three days of hospitalization [6], when prognosis is most uncertain. A new treatment that promotes early recovery of consciousness in the ICU would benefit patients and families by reducing the likelihood of premature withdrawal of life-sustaining therapy and may decrease ICU complications related to immobility, facilitate self-expression, enable autonomous decision-making, and increase access to specialized rehabilitative care.

Two barriers currently prevent the development of consciousness-promoting therapies in the ICU. First, patients are not rigorously classified prior to enrollment in clinical trials. Instead of selecting patients based on the precise pathophysiologic mechanism underlying coma, trials enroll patients based on behavioral measures, such as the Glasgow Coma Scale (GCS) score, that divide brain injuries into severity categories. This approach is ineffective because coma is a highly heterogeneous condition. Traumatic and hypoxic-ischemic coma, for example, are associated with variable patterns of axonal disconnections within the subcortical ascending arousal network (AAN), cortical default mode network (DMN), and other networks that contribute to consciousness [9-14]. Without precision tools to map preserved network connections in individual patients, it is not possible to identify patients whose connectomes are amenable to therapeutic modulation.

Second, therapeutic responses are not routinely tested with direct measures of brain function in early-phase clinical trials. Rather, early-phase trials rely on indirect serologic markers of brain injury or insensitive, delayed measures of functional disability [15, 16]. Without biomarkers that directly and quantitatively measure brain function, fundamental questions about a therapy’s neurobiological effects are not being answered in Phases 1 and 2 before moving to Phase 3 trials. Clearly, a new approach to clinical trial design is of scientific and ethical import for this vulnerable population [5]. Indeed, experts in civilian and military brain injury [15-17], including leaders at the National Institute of Neurological Disorders and Stroke [18], are now calling for new approaches to clinical trial design for comatose patients. This goal is also a central component of the Curing Coma Campaign launched by the Neurocritical Care Society in 2019.

To address the call for clinical trial design innovation, we propose the <u>C</u>onnectome-based <u>C</u>linical <u>T</u>rial <u>P</u>latform (CCTP), a new mechanistic paradigm for developing and testing targeted therapies that promote early recovery of consciousness in the ICU. The CCTP incorporates two innovations: 1) predictive biomarkers to enroll patients in clinical trials based on connectomes that can be targeted by new therapies; and 2) pharmacodynamic biomarkers to measure how targeted therapies modulate networks, reactivate the cerebral cortex and restore consciousness. By enrolling patients in clinical trials based on a principled, mechanistic assessment of their neuroanatomic potential for a therapeutic response, the CCTP will generate enhanced study samples of patients with similar connectomes, thereby increasing treatment effect sizes, decreasing sample sizes needed to power trials, and ultimately reducing trial duration, cost and risk to subjects [16]. Most importantly, by enhancing the scientific rigor of early-phase clinical trials, we hypothesize that the CCTP will improve the success rate of late-phase trials so that investigators can bring new therapies to clinical practice in the ICU. To demonstrate the utility of implementing the CCTP in the ICU, we describe STIMPACT (<u>S</u>timulant <u>T</u>herapy Targeted to Individualized Connectivity <u>M</u>aps to <u>P</u>romote Re<u>ACT</u>ivation of Consciousness), a CCTP-based clinical trial in which intravenous methylphenidate (IV MPH) will be used to promote recovery of consciousness in severely brain-injured patients in the ICU (ClinicalTrials.gov NCT03814356).

## Methods

### Conceptual and Empiric Basis for the CCTP

Current theories propose that consciousness requires the integrated function of multiple brain networks, not any single node or connection [19, 20]. This network-based model of consciousness is supported by histologic and radiologic data demonstrating that coma can be caused by variable network disconnections [9, 10, 21-23], and that consciousness requires dynamic network interactions [24]. For a patient to recover consciousness, it is believed that the subcortical AAN, which modulates wakefulness [25, 26], must reconnect with the DMN and other cortical networks, which mediate awareness [13, 27]. However, the precise mechanisms underlying this reconnection are poorly understood. Furthermore, it may be weeks, months, or even years before these recovery mechanisms allow preserved AAN connections to reactivate the cerebral cortex [28-30]. For patients whose families face time-sensitive, life-or-death decisions in the ICU, treatments that promote recovery are needed within days. The CCTP will accelerate recovery of consciousness via targeted therapeutic modulation of structurally preserved brain network connections. The ultimate goal of the CCTP is to provide ICU clinicians with an armamentarium of targeted therapies that promote recovery of consciousness in patients with a broad range of coma etiologies.

### Testing the CCTP – Patient and Target Selection for STIMPACT

STIMPACT will test the CCTP in patients with acute severe TBI who are being treated in the ICU. We selected this patient population for our inaugural trial because prior evidence suggests that components of the AAN may be spared, and therefore serve as therapeutic targets, in patients with severe TBI. Histopathological studies show that although the AAN is invariably injured in animals and humans with coma caused by severe TBI [10, 31], not all AAN nuclei are lesioned and not all axons are disconnected. For example, both histopathological and MRI studies reveal that the ventral midbrain, home to the dopaminergic ventral tegmental area (VTA), often remains at least partially intact after severe TBI [22, 32-34]. This phenomenon pertains not only to VTA neuronal cell bodies in the midbrain, but also to VTA axons that connect with the diencephalon, basal forebrain, and cortex (Figures 1, 2). Consistent with these observations, nuclear imaging studies show that signal transmission in dopaminergic circuits is variably altered by severe TBI [35-37]. These converging lines of evidence support the hypothesis that preserved VTA connections are a potential target for therapeutic modulation in ICU patients with severe TBI.

**Figure 1:**
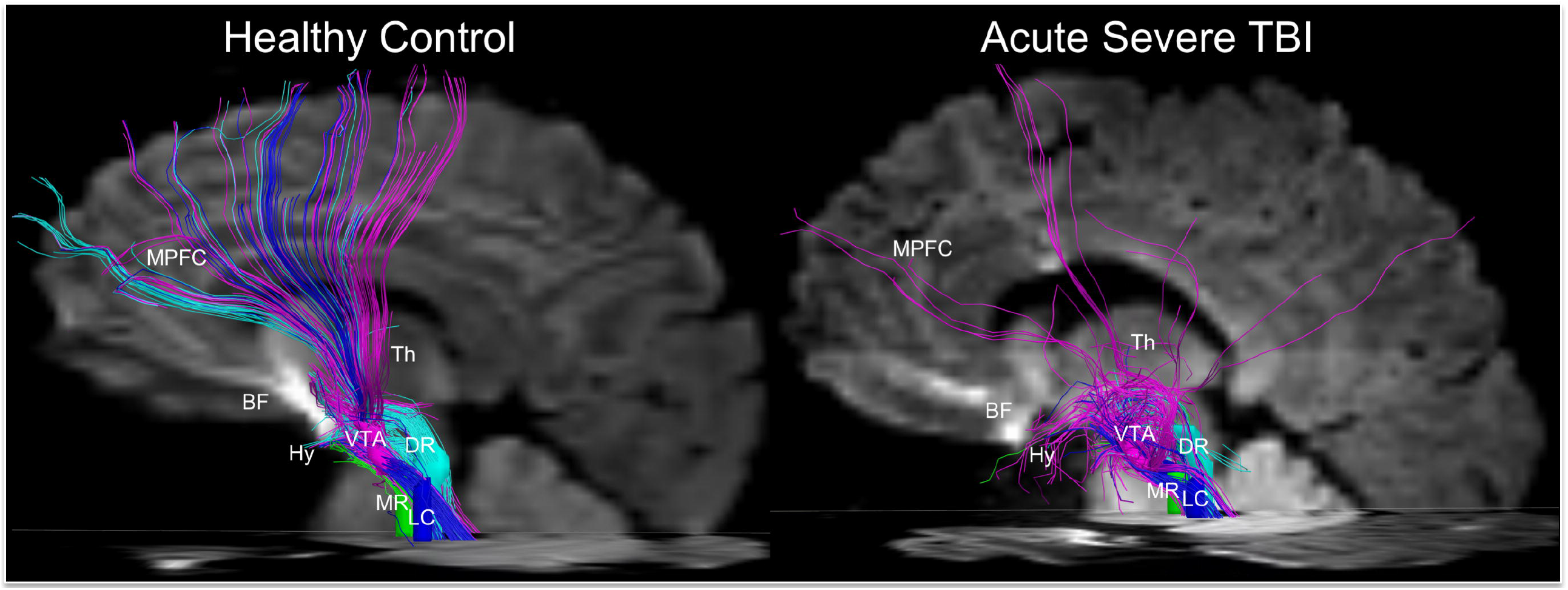
Personalized Connectome Mapping in the ICU Reveals Preserved Ventral Tegmental Area Connections. VTA tracts are shown from a left lateral view in a 33-year-old healthy male control and a 29-year-old man with acute severe TBI. The patient was comatose on arrival to the hospital and in a minimally conscious state at the time of this scan on post-injury day 7, as determined by a Coma Recovery Scale-Revised assessment. Tractography analysis was performed using TrackVis, as previously described [26]. All tracts are color-coded by sites of VTA connectivity: turquoise with dorsal raphe (DR), blue with locus coeruleus (LC), green with median raphe (MR), and pink with cortex, thalamus (Th), hypothalamus (Hy), or basal forebrain (BF). Multiple VTA connections are preserved in the patient, including with the medial prefrontal cortex (MPFC), which is a node of the default mode network. The patient recovered consciousness and functional independence by 6 months. Even in a patient with acute severe traumatic brain injury, the VTA may be a hub through which multiple brainstem AAN nodes connect with the Th, Hy and cerebral cortex.

**Figure 2:**
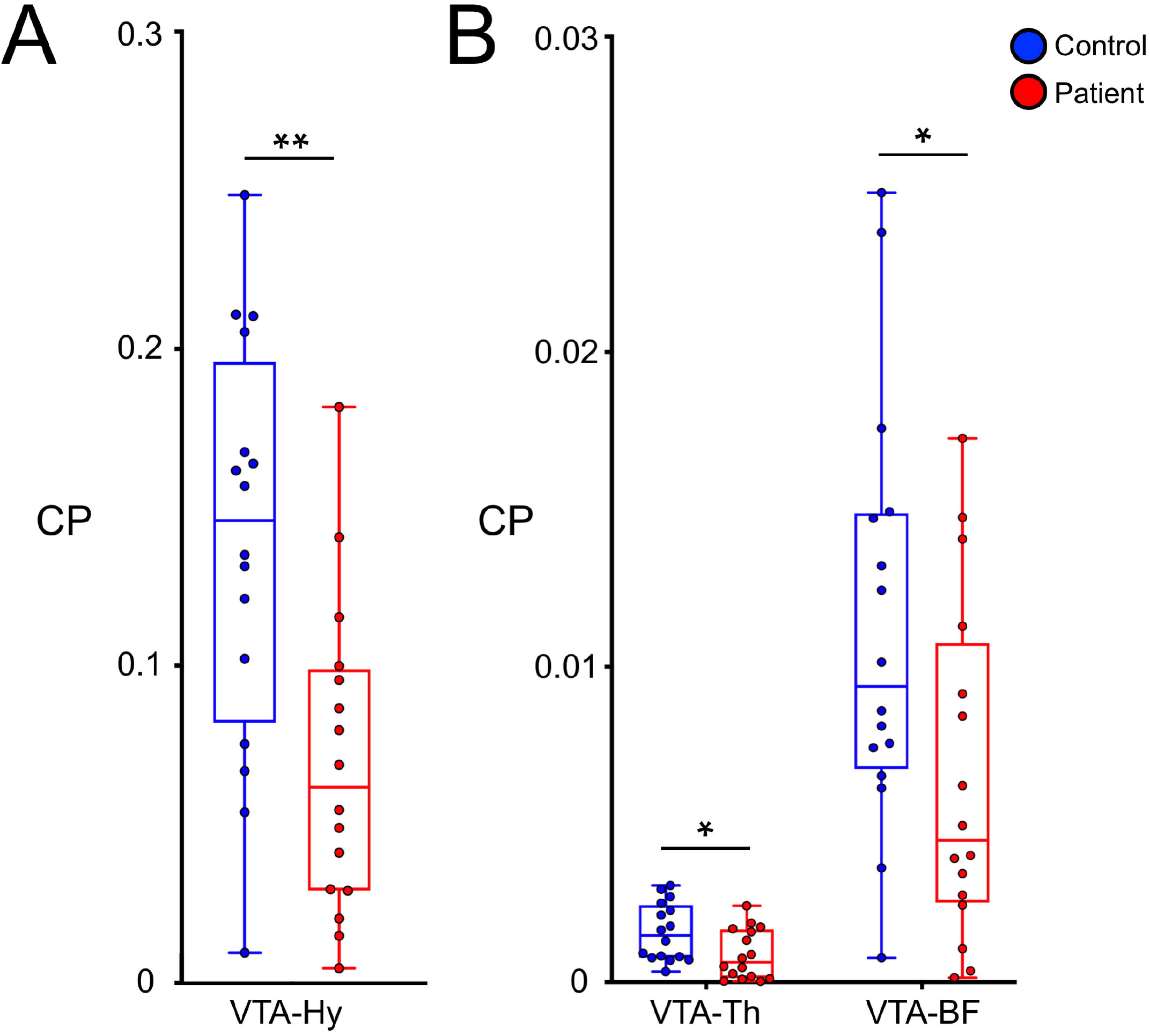
VTA Axonal Connections are Spared in a Subset of Patients. We analyzed VTA connectivity with the hypothalamus (Hy), thalamus (Th), and basal forebrain (BF) in 16 patients, as well as in 16 matched controls. We calculated a connectivity probability (CP) between the VTA and each target AAN node using previously described methods [47, 71]. On a group level, median CP values were lower between the VTA and each AAN node in patients compared with controls. However, there is significant variance in control-group CP values, and many patients fall within the control-group interquartile range (IQR). These results indicate relative sparing of VTA connections in a subset of patients with acute severe TBI. See the Supplementary Materials for a detailed description of the CP calculation algorithm.

### Testing the CCTP – Therapy Selection for STIMPACT

We selected IV MPH as the therapeutic agent for STIMPACT because of its well-established mechanism of action on the VTA, compelling rodent data suggesting its role in promoting consciousness, and decades of data indicating its safe use in humans. IV MPH is a dopamine reuptake inhibitor that potentiates dopaminergic neurotransmission by VTA neurons [38, 39]. Although multiple neurotransmitters contribute to arousal [25], empirical evidence strongly supports the role of dopamine in therapeutic modulation of consciousness.

In rodent models of anesthetic coma, IV MPH promotes reemergence of consciousness by stimulating dopaminergic VTA neuronal signaling via D1 receptors [38, 40]. Furthermore, optogenetic [41] and electrical stimulation [40] of dopaminergic VTA neurons promotes recovery of consciousness in anesthetized rodents. In humans, multiple enteral and subcutaneous therapies that stimulate dopaminergic neurotransmission have been shown to benefit patients with subacute or chronic TBI, including enteral MPH [42, 43]. Moreover, enteral amantadine, the only therapy shown to promote recovery of consciousness in a randomized controlled trial of patients with subacute TBI [44], increases dopamine release and blocks dopamine reuptake, thereby increasing synaptic levels of dopamine. Most prior trials investigated enteral MPH, which is FDA-approved to treat attention deficit disorder [39] and prescribed “off-label” to promote recovery in patients with TBI [45, 46]. However, positron emission tomography studies show that the IV formulation of MPH has a far more rapid uptake in the human brain than does the enteral formulation (∼10 minutes versus ∼60 minutes) [39]. Collectively, prior animal and human data provide a biological basis for using IV MPH to upregulate structurally intact but functionally dormant dopaminergic VTA circuits in ICU patients with severe TBI.

### STIMPACT Hypothesis

The central hypothesis of the STIMPACT trial is that preservation of VTA connections within the AAN connectome is a predictive biomarker of a response to IV MPH in patients with acute severe TBI. We hypothesize that for the subset of patients with preserved VTA connections, IV MPH will reconnect the cerebral cortex, accelerate reemergence of consciousness, and transform the course of their recovery.

### Predictive Biomarker for STIMPACT

To identify patients with preserved VTA connections, we will map the structural AAN connectome on a clinical MRI scanner. We will use high angular resolution diffusion imaging (HARDI) tractography, an MRI technique that maps axonal connectivity based on directional water diffusion [47]. We have previously shown that HARDI tractography methods can map the AAN connectome in the *ex vivo* human brain [26], detect acute AAN disconnections *ex vivo* [10], and detect disruptions *in vivo* in ICU patients with acute severe TBI [9].

Given that the VTA and cerebral cortex are connected both monosynaptically and polysynaptically [26, 48] (Figure 3), it is unlikely that a single preserved VTA connection will predict a patient’s response to IV MPH. Rather, we will use a graph theoretical analysis measure, VTA hub strength (*S*_*VTA*_), as a predictive biomarker for the STIMPACT trial, because this biomarker measures both direct (monosynaptic) and indirect (polysynaptic) VTA connections. We selected subcortical AAN and cortical DMN nodes as targets in the *S*_*VTA*_ measurement because functional connectivity studies suggest that DMN reactivation is essential for recovery of consciousness after severe brain injury [12, 49-52] and that the VTA activates the DMN via the AAN [53].

**Figure 3:**
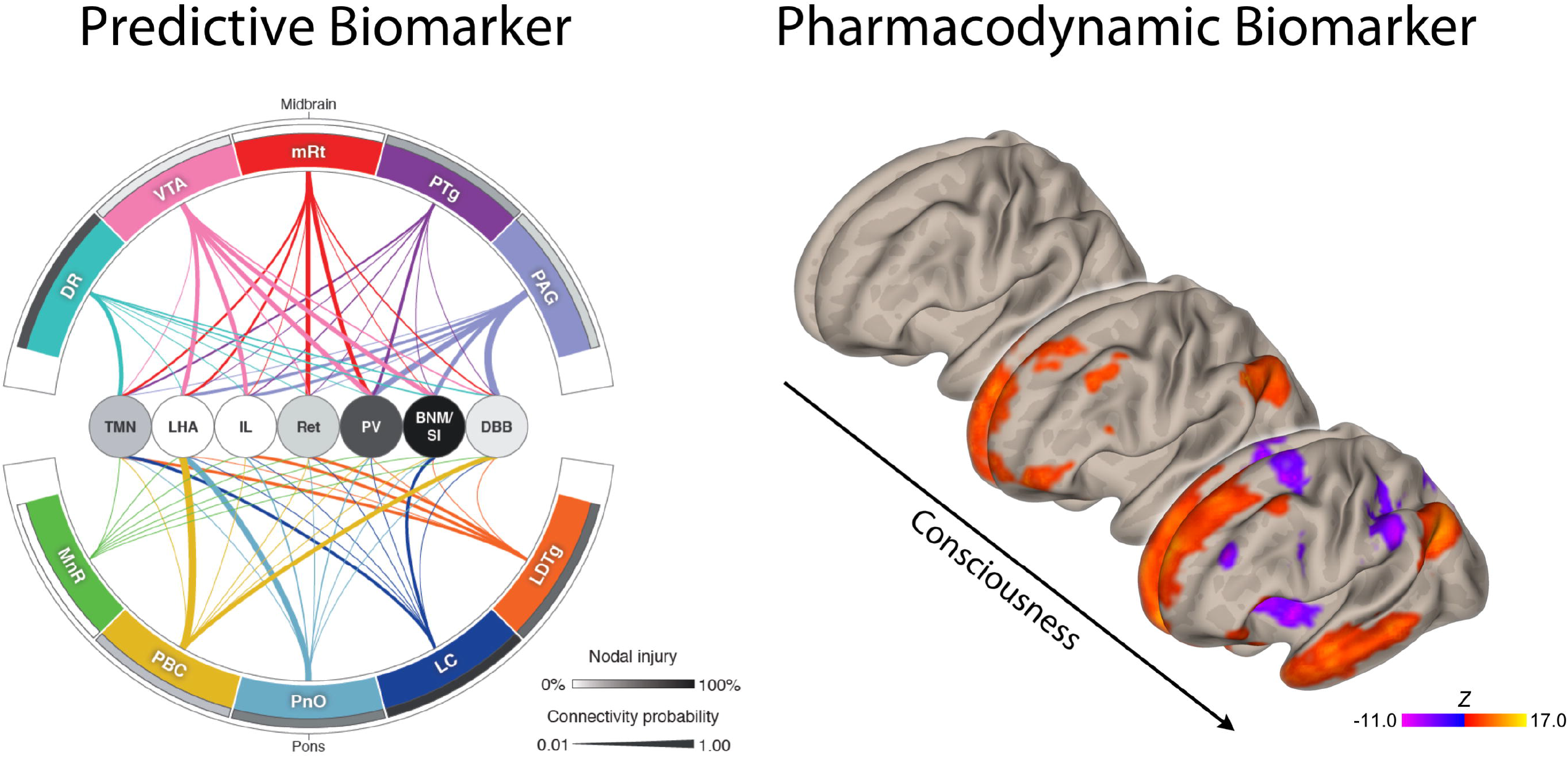
Predictive and Pharmacodynamic Biomarkers. Panel A: This type of individualized ascending arousal network (AAN) connectome map will be used as a biomarker to predict patient responses to therapy. In this sample “connectogram,” brainstem nodes are shown on the outside, while hypothalamic, thalamic, and basal forebrain nodes are shown in the middle. Line thickness is proportional to the connectivity probability (CP; see Supplementary Materials for how this value is measured) for each node-node pair. Nodal grey shading is proportional to the percentage of each node occupied by a traumatic lesion (bottom right bar). These structural connectivity data, along with structural connectivity measures between the VTA and default mode network (DMN), are used to calculate *S*_*VTA*_. Connectogram artwork by Kimberly Main Knoper. Panel B: Resting-state functional MRI maps illustrating concurrent recovery of consciousness and reactivation of the DMN. Hot colors indicate correlated activity within the DMN. Cool colors indicate regions anti-correlated with the DMN (inset). Functional connectivity between the VTA and DMN (Z_*VTA-DMN*_) is a rs-fMRI pharmacodynamic biomarker that will be used to determine the neurobiological effects of intravenous methylphenidate in patients with acute severe traumatic brain injury. Abbreviations: nucleus basalis of Meynert/substantia innominata (BNM/SI), diagonal band of Broca (DBB), dorsal raphe (DR), intralaminar nuclei of the thalamus (IL), lateral hypothalamic area (LHA), laterodorsal tegmental nucleus (LDTg), locus coeruleus (LC), median raphe (MnR), mesencephalic reticular formation (mRt), parabrachial complex (PBC), paraventricular nucleus of the thalamus (PV), pedunculotegmental nucleus (PTg), periaqueductal grey (PAG), pontis oralis (PnO), reticular nuclei of the thalamus (Ret), supramammillary nucleus of the hypothalamus (SMN), tuberomammillary nucleus of the hypothalamus (TMN), ventral tegmental area (VTA).

### Pharmacodynamic Biomarkers for STIMPACT

To measure the pharmacodynamic effects of IV MPH, we will map the AAN’s functional connectome with a multimodal approach that integrates resting-state functional MRI (rs-fMRI) and EEG. First, we will use blood-oxygen-level dependent (BOLD) rs-fMRI to generate Fisher Z-normalized measurements of VTA-DMN functional connectivity (*Z*_*VTA- DMN*_). We and others have demonstrated the feasibility of rs-fMRI mapping of functional brain networks in ICU patients with acute severe TBI (Figure 3) [12, 50, 51], and in a preliminary analysis we observed that *Z*_*VTA-DMN*_ increases as patients recover consciousness (Figure 4). Given that brainstem functional network mapping requires high spatial and temporal resolution, we implemented a simultaneous multislice [54] BOLD sequence on the 3 Tesla Siemens Skyra MRI scanner in the Massachusetts General Hospital Neurosciences ICU (2 mm isotropic voxels, TR=1.25 seconds; see Supplementary Material for additional sequence parameters). To control for the effects of physiological fluctuations on the BOLD signal [55], we measure and statistically regress cardiac and respiratory activity during rs-fMRI (Figure 5 and Supplementary Figures 1 and 2). Details regarding physiologic data acquisition and analysis are provided in the Supplementary Methods.

**Figure 4:**
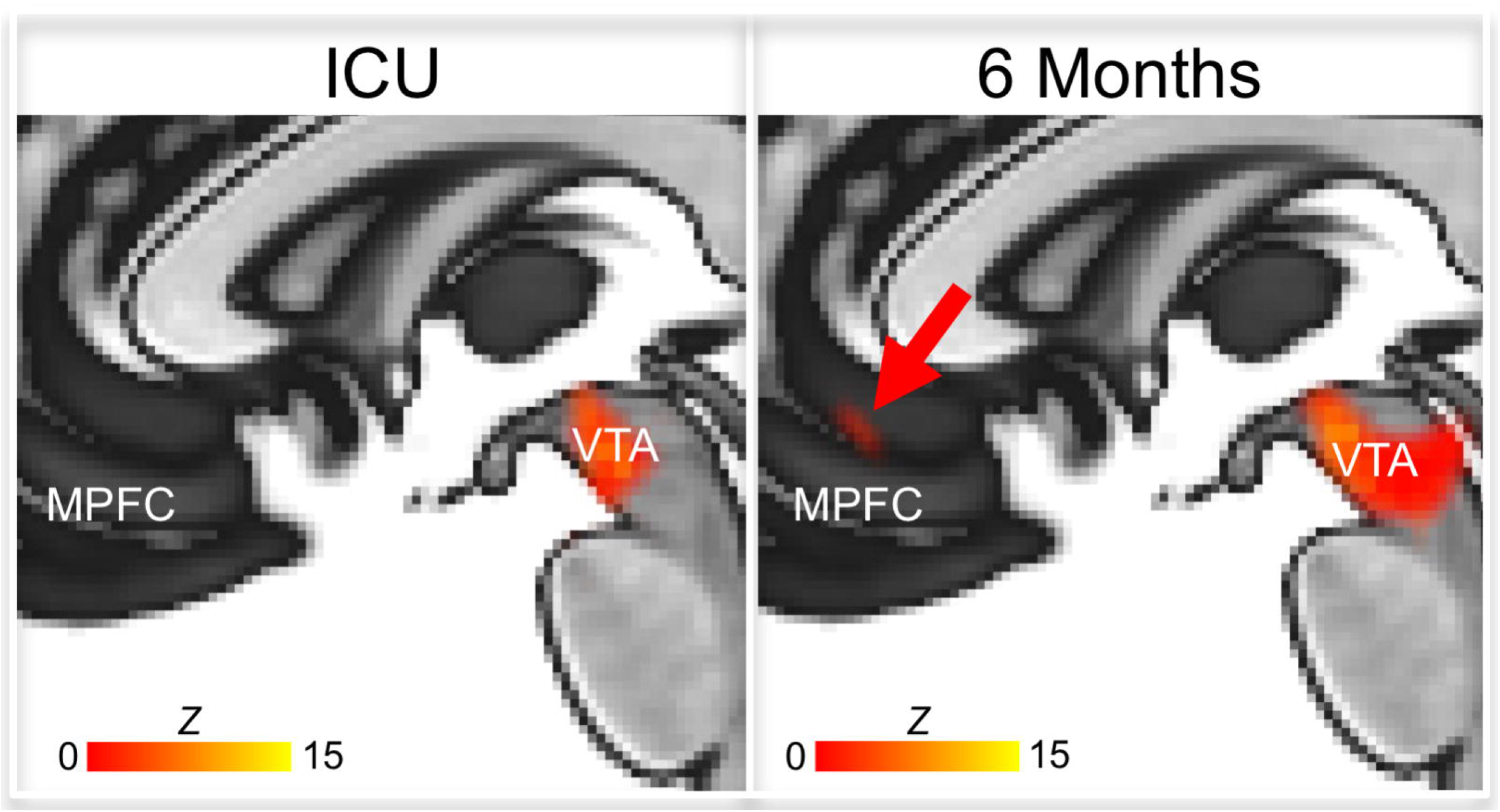
Functional Connectivity between the Ventral Tegmental Area and Default Mode Network Reemerges During Recovery. Eight patients with acute severe TBI underwent rs-fMRI in the ICU and at six-month follow-up, by which time all had recovered consciousness. Mean group-level VTA functional connectivity maps are shown, based on Fisher Z-transformed correlations in the BOLD signal (color bar). There were no VTA-DMN connections acutely, but VTA connections with the medial prefrontal cortex (MPFC) node of the DMN reemerged during recovery of consciousness (red arrow). The statistical threshold used here is more liberal than in our recent studies of DMN connectivity [12, 50], which underscores the need for the higher-resolution rs-fMRI sequence that we developed for the CCTP.

**Figure 5:**
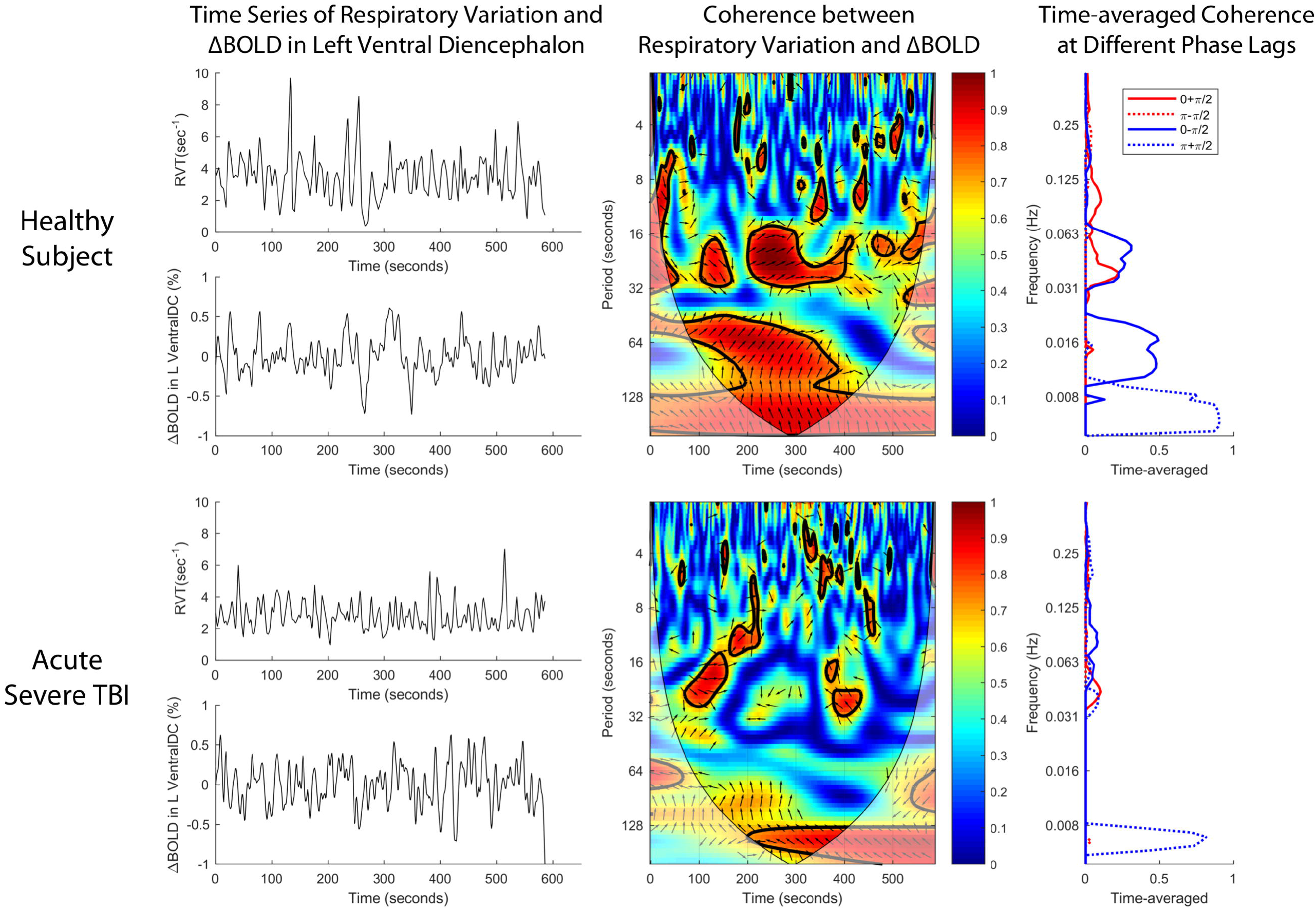
Wavelet Transform Coherence Analysis of a Representative Control Subject and a Representative Patient. Time series of respiratory variation and change in blood-oxygen level dependent (ΔBOLD) signal in the left ventral diencephalon are shown in the left column. The dynamic interaction between respiratory variation and ΔBOLD is demonstrated by the squared wavelet coherence map between the time series of respiratory variation and ΔBOLD shown in the middle column. The magnitude of coherence ranges between 0 and 1, where warmer color represents stronger coherence and cooler color represents weaker coherence. Significant coherence between respiratory variation and ΔBOLD occurs in the area defined by the thick contour of the unfaded region. The x-coordinate of the area provides information on the duration of the oscillating cycle when respiratory variation interacts with ΔBOLD, and the y-coordinate shows the time when this interaction occurs over the resting state fMRI scan. The simplified format of coherence between respiratory variation and ΔBOLD is shown in the right column, with the features of oscillations displayed in terms of frequency. While increased coherence is found between respiratory variation and ΔBOLD at the frequency range of 0.008-0.063Hz in the healthy subject, the coherence between respiratory variation and ΔBOLD in the same frequency range is diminished in the patient with acute severe TBI. Compared with the healthy subject, the resting state BOLD signal changes have less influence from respiratory variation in the patient with acute severe TBI. A detailed interpretation of the wavelet coherence findings is included in Supplementary Materials.

Second, we will use resting-state EEG data to identify a quantitative EEG biomarker that measures the effect of IV MPH on cortical circuit function. Crucial to this effort is the development of statistical methods that quantify EEG oscillatory dynamics. Patients with DoC experience frequent fluctuations in arousal [56], and the dynamic range and robustness of EEG biomarkers during these fluctuations in the ICU environment is unknown. Therefore, we will acquire 24 hours of baseline EEG data for each patient prior to the first IV MPH dose, allowing therapeutic responses to be compared to each patient’s baseline biomarker variance.

Candidate EEG biomarkers include quantitative measures from spectral [57], graph theoretical [58], complexity [59], and entropy [60] analyses. Efforts to compare the diagnostic utility of these biomarkers are underway in animals [61], patients with chronic DoC [62], and ICU patients with acute DoC [63-65]. Recent studies of patients with chronic DoC suggest that spectral measures perform well in comparison to other EEG-based cortical measures [62], and our preliminary results suggest that patients experience wide-ranging fluctuations in frequency band-specific activity over the course of a clinical recording (Figure 6). Nevertheless, the ratio of alpha to delta power appears to reliably distinguish patients with DoC from patients who have recovered consciousness. Ongoing efforts are underway to determine which EEG biomarkers are most responsive to therapeutic stimulation in patients with acute DoC.

**Figure 6:**
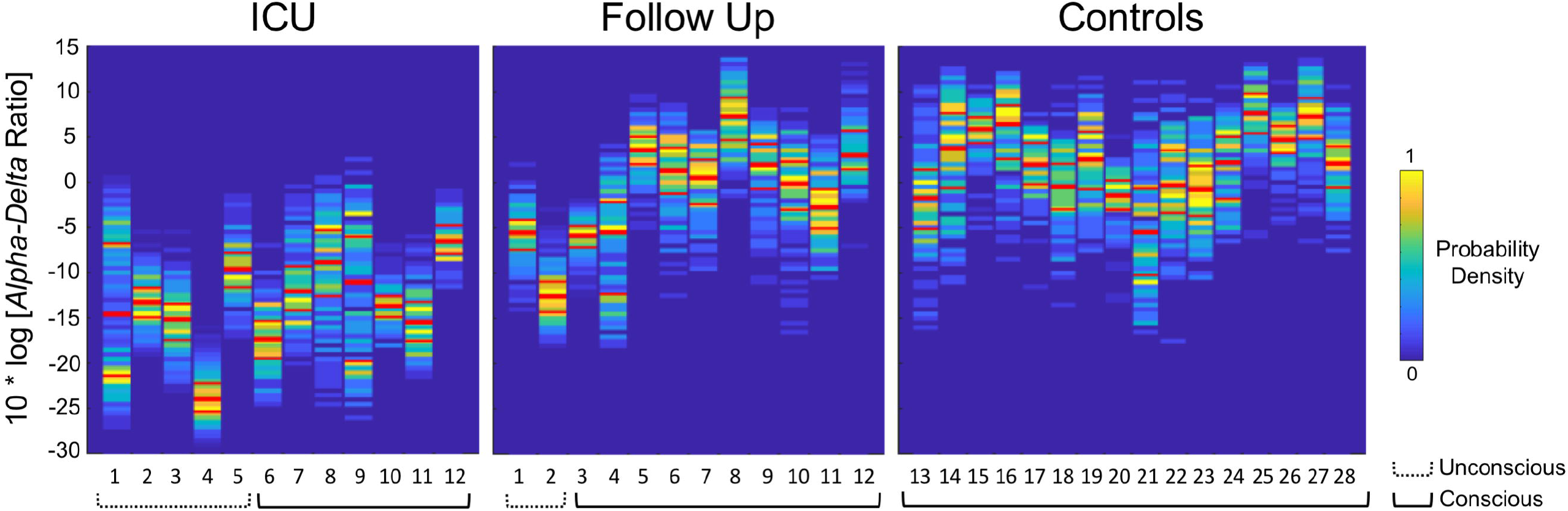
Alpha-delta Ratio as a Biomarker of Recovery of Consciousness. We computed the dynamic range of the resting-state *alpha-delta* ratio in EEG recordings of patients with severe traumatic brain injury and controls. The plots show comparisons of *alpha-delta* ratio measures in 12 ICU patients with acute severe traumatic brain injury (left), these same 12 patients at 6-month follow-up (middle), and 16 healthy controls (right). Each column represents a single subject. The three red lines within each subject’s *alpha-delta* ratio plot represent the interquartile range (outer red lines) and the median (middle red line). These results suggest that 1) *alpha-delta* ratio is generally lower in unconscious ICU patients (coma and vegetative state) than in conscious ICU patients (minimally conscious state and post-traumatic confusional state); 2) in ICU patients who recover consciousness by 6 months, *alpha-delta* ratio typically increases to values similar to those of controls; and 3) there is substantial intra-subject variance in *alpha-delta* ratio during a single EEG recording.

### Safety Considerations and Rationale for a Phase 1 Trial

No serious adverse events (SAEs) were reported in the 1,700 humans who received IV MPH between 1958 and 2010. Common drug-related side effects include hypertension, tachycardia, agitation, and nausea [66], which are readily manageable in the ICU environment of the STIMPACT trial. However, IV MPH has never been administered to a patient with acute severe TBI, and experience with IV MPH in comatose humans is limited to small case series of patients with barbiturate overdoses [67-69]. Thus, we believe that it is scientifically and ethically appropriate to begin STIMPACT with a Phase 1 safety study.

Notably, there is a hypothetical risk that early stimulant therapy in the ICU could harm patients by exacerbating glutamate-mediated excitotoxicity in the traumatized brain. Yet, to our knowledge, there is no evidence from animal or human studies to substantiate this risk. Rather, evidence from a rat neuronal culture study suggests that dopamine may be protective against excitotoxicity [70] and we recently found that enteral stimulants, including MPH, were well tolerated with no SAEs in 48 ICU patients [45].

### Overview of STIMPACT Phase 1 Study Design

In Phase 1 of STIMPACT, we aim to enroll 22 adult patients with acute traumatic DoC regardless of *S*_*VTA*_ magnitude. Inclusion and exclusion criteria are described on clinicaltrials.gov (NCT03814356). Because patients with acute severe TBI have increased hepatic drug metabolism and clearance, we cannot rely on prior dose-finding and pharmacokinetic studies of healthy humans [39, 66] and must determine the maximum tolerated dose and pharmacokinetics of IV MPH in our population of interest. Thus, our primary endpoint is the number of drug-related SAEs occurring at each of three IV MPH doses: 0.5, 1.0 and 2.0 mg/kg. Our secondary endpoint is the effect of IV MPH on *Z*_*VTA-DMN*_ and the *alpha-delta* ratio. These rs-fMRI and EEG metrics will provide a direct pharmacodynamic assessment of IV MPH on its target brain network.

Each patient will undergo 5 days of data acquisition, with predictive biomarker data and baseline pharmacodynamic biomarker data collected on Day 0, and treatment-related biomarker data collected on Days 1-4 (Figure 7). Hence, each patient’s biomarker responses to IV MPH will be measured against individual baseline biomarker variance – this will help account for each patient’s current drug regimen, which will be determined by clinical need. IV MPH will be administered as daily boluses (via IV push at a rate not to exceed 20 mg/min) on Days 1-4: 0.5 mg/kg on Day 1, 1.0 mg/kg on Day 2, 2.0 mg/kg on Day 3, and the maximum tolerated dose on Day 4. On Days 1-3, we will record EEG continuously to measure *alpha-delta* ratio before and after each dose. On Day 4, each patient will receive his/her maximum tolerated dose during an rs-fMRI scan to measure *Z*_*VTA-DMN*_ prior to and after IV MPH.

**Figure 7:**
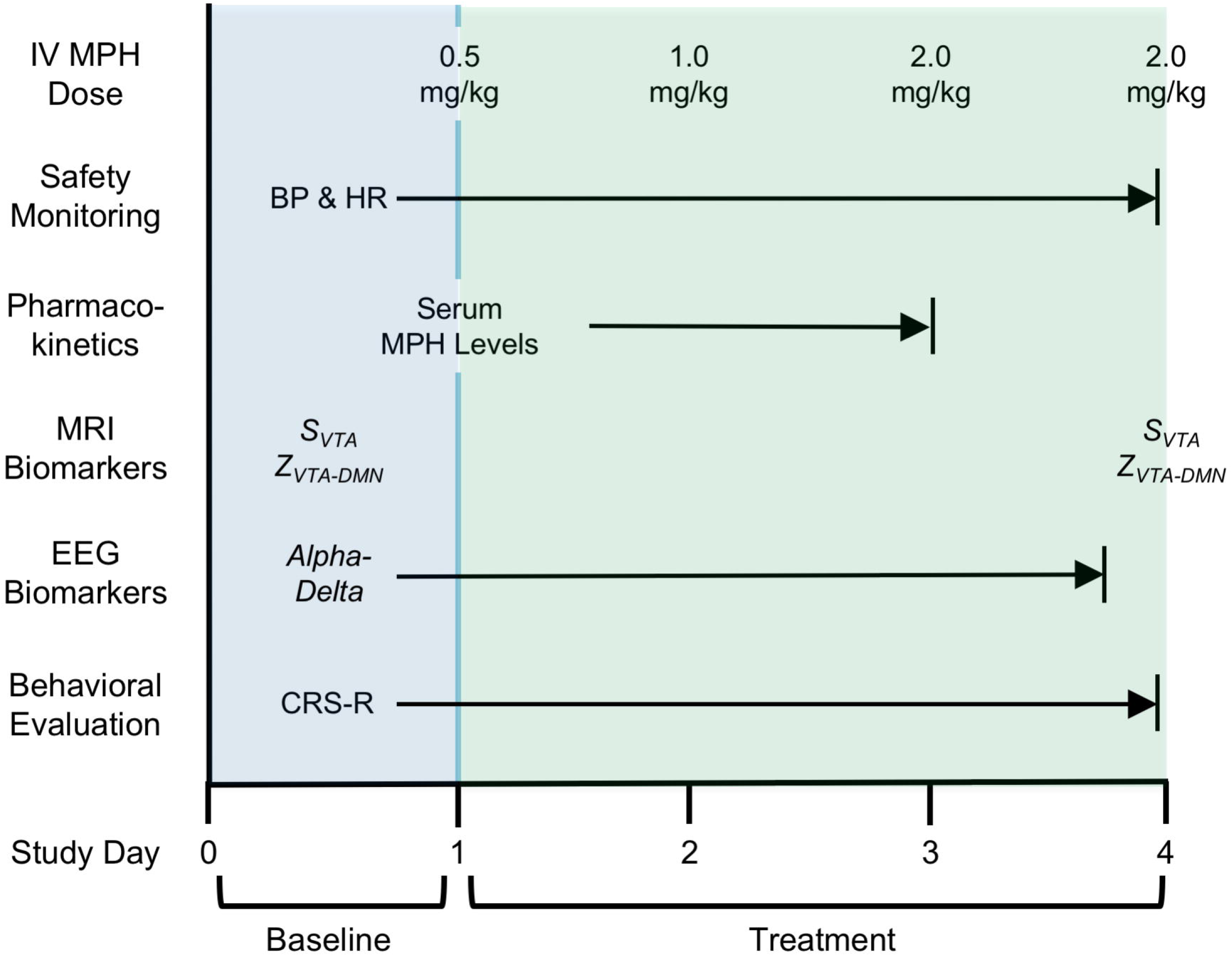
STIMPACT Study Design Schematic. Each patient in Phase 1 of the STIMPACT Trial will undergo five days of data acquisition, with predictive biomarker data and baseline pharmacodynamic biomarker data collected on Day 0, and treatment-related biomarker data collected on Days 1-4. Hence, each patient’s biomarker responses to IV MPH will be measured against his/her own baseline biomarker variance. Abbreviations: BP = blood pressure; CRS-R = Coma Recovery Scale-Revised; HR = heart rate; MPH = intravenous methylphenidate; *S*_*VTA*_ = ventral tegmental area hub strength; *Z*_*VTA-DMN*_ = functional connectivity between the ventral tegmental area and the default mode network.

While broader than the 0.5 to 0.9 mg/kg dose range used in most prior human studies [39, 66], the range of 0.5 to 2.0 mg/kg may still be insufficient to detect a pharmacodynamic response in patients with severe TBI. Therefore, we require a pharmacodynamic response in ≥10% of patients in Phase 1 to proceed to Phase 2, a double-blind, placebo-controlled, cross-over trial which will test the hypothesis that preserved *S*_*VTA*_ predicts a pharmacodynamic biomarker response to IV MPH relative to placebo. Without such a response, we will repeat Phase 1 at a higher dose range. A detailed statistical analysis plan for the STIMPACT Phase 1 trial is provided in the Supplementary Material.

In July 2018, we convened a Clinical Oversight Committee, led by an experienced Independent Medical Monitor (Dr. Thomas P. Bleck), which approved our approach to conducting the STIMPACT trial. To ensure that the TBI community’s interests are represented, we also convened a Patient and Family Advisory Board, which reviewed the research plan and made recommendations regarding recruitment, study activities, and dissemination. The investigational new drug (IND) application was approved by the FDA in August, 2018 (IND# 140675), and the trial protocol was approved by the Partners Healthcare Institutional Review Board in May, 2019.

## Summary

We propose a mechanistic clinical trial paradigm, the Connectome-based Clinical Trial Platform (CCTP), to predict and measure responses to new therapies in the ICU. We also describe a CCTP-based trial, STIMPACT, which will test the hypothesis that IV MPH promotes recovery of consciousness in patients who have preserved VTA connections within their AAN connectomes. The CCTP will allow clinicians to provide targeted treatments that are personalized to each patient’s connectome, ensuring that each patient is given the best possible chance to recover consciousness in the ICU. Ultimately, we envision multidisciplinary discussions about optimal treatment selection at a *Coma Board*, akin to the *Tumor Board* at which therapeutic decisions are made by a multidisciplinary team for patients with cancer. By providing a clinical trial platform to test targeted pharmacologic and electrophysiologic therapies that reactivate the injured human connectome, the CCTP has the potential to transform the therapeutic landscape and improve outcomes for patients with coma.

## Data Availability

Data processing scripts can be found at: github.com/ComaRecoveryLab/AAN-in-Acute-Traumatic-Disorders-of-Consciousness.git. The conditions of our Institutional Review Board ethics approval do not permit public archiving of anonymized study data. Readers seeking access to the data should contact the corresponding author.

## Acknowledgments

We thank the members of the Patient and Family Advisory Board of the Massachusetts General Hospital Laboratory for NeuroImaging of Coma and Consciousness for their feedback and insights regarding the ethical conduct of this clinical trial. We also thank Dr. David A. Schoenfeld for helpful statistical consultation.

